# On the secondary waves of the pandemic launched in Iran and other countries

**DOI:** 10.1101/2020.06.20.20136283

**Authors:** D. Kovriguine, S. Nikitenkova

## Abstract

In the last decade of April 2020, the second epidemic wave began in Iran. If judging by the dynamics of total cases, the birth of this second wave coincides practically with the maximum growth rate of the primary epidemic started in Iran from the very beginning of 2020. Today, the secondary epidemic wave almost doubles the peak of the primary. The new epidemic wave grows rapidly and unpredictably. Also, we admit that the secondary epidemic waves are already getting started in other countries.

## Introduction

The first wave of the epidemic in Iran has attracted a permanent interest of observers and researchers because of relatively sluggish volatility accompanied a lot of total cases [1]–[5]. Recently, we have started to suspect that the first wave was only a harbinger of the spread of a new, higher wave of the epidemic which is now rapidly spreading throughout this country.

The secondary waves of the epidemic are also visible in some other countries. In Sweden, the secondary epidemic wave has overcome the peak of the primary almost three times, for example. A cascade of numerous secondary waves of relatively low intensity, but recurring with enviable regularity, takes place in the United States.

The purpose of this work is to mathematically describe the second wave of the epidemic in Iran using a simple dynamic system. We have taken the logistic curve as the predicted result since it requires a minimum of information for identification. We need to know are the initial condition, the peak value, and the number of days from the start to the epidemic peak.

## Method

To build a dynamic system, we use the tabular autocorrelation dependencies *N*_*i+*1_ = *f*_*c*_ (*N*_*i*_). Here, *N*_*i*_ is the number of total cases, accumulated to the date *i*, in the country number *c*. Using the least-squares method, we determine the polynomial approximations of tabular functions *f*_*c*_, denoted as *F*_*c*_. Numerical experiments confirm the sufficiency of the quadratic approximation of functions *f*_*s*_ from the argument *N*_*i*_.

So, the dynamic model takes the form of an analytic point mapping with the right-hand side 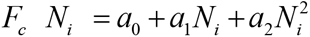. Here, *a*_*i*_ are the parameters determined from the tables [6]. The transition time between the states *N*_*i*_ and *N*_*i +*1_ is a day. After processing the data, we select results using simple criteria. In the case of fuzzy logic, the user of the algorithm can verify and interpret the obtained results.

We suppose that the logistic equation describes the temporal evolution of each wave of the epidemic adequately in countries with high-quality statistics. The phase point of the dynamic system moves from the unstable state *N =* 0 to the stable steady-state *N =N*^*^. Here, *N*^*^ is the positive root of the quadratic equation *N* ^*^ = *a*_0_ *+ a*_1_ *N* ^*^ +*a*_2_ *N* ^*2^. The steady-state *N*^*^ corresponds to almost zero dynamic increase in disease. At the time of achieving this stable equilibrium, the logistic equation is no longer sufficient to describe the evolution of the disease in a developed human society. The fate of infected people depends on the quality of quarantine measures, medical cares, etc.

There is no doubt that, theoretically, the epidemic can develop for the second and third time, etc. in an unpredictable way, if we neglect the appropriate anti-epidemic measures. As calculations show, the second wave of the epidemic, as a rule, rises after passing the threshold point, which is the inflexion point of the logistic curve. The linear equation 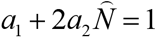 defines the threshold 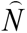. In the emergency of a second epidemic wave, we use the same dynamic system for calculations. Following a simple algorithm, we calculate the new parameters *a*_*i*_ using the new input data. We remove the already calculated data, provided by the logistic curve of the first wave, from the old input data set. The computation process continues until the input data will represent a superposition of various individual logistic curves. There is no doubt that all input is perfect. In case of failure of the specified decomposition of the input data, we decide that the algorithm described above does not work, and omit such input data from the analysis.

Now, let us consider an example of the epidemic history in a country that already has reached the peak of the first wave, and gave rise for the second wave. Let it be the most vividly manifested country in this respect, Iran, whose the epidemic history presented in Fig. 1.

**Fig. 1.**
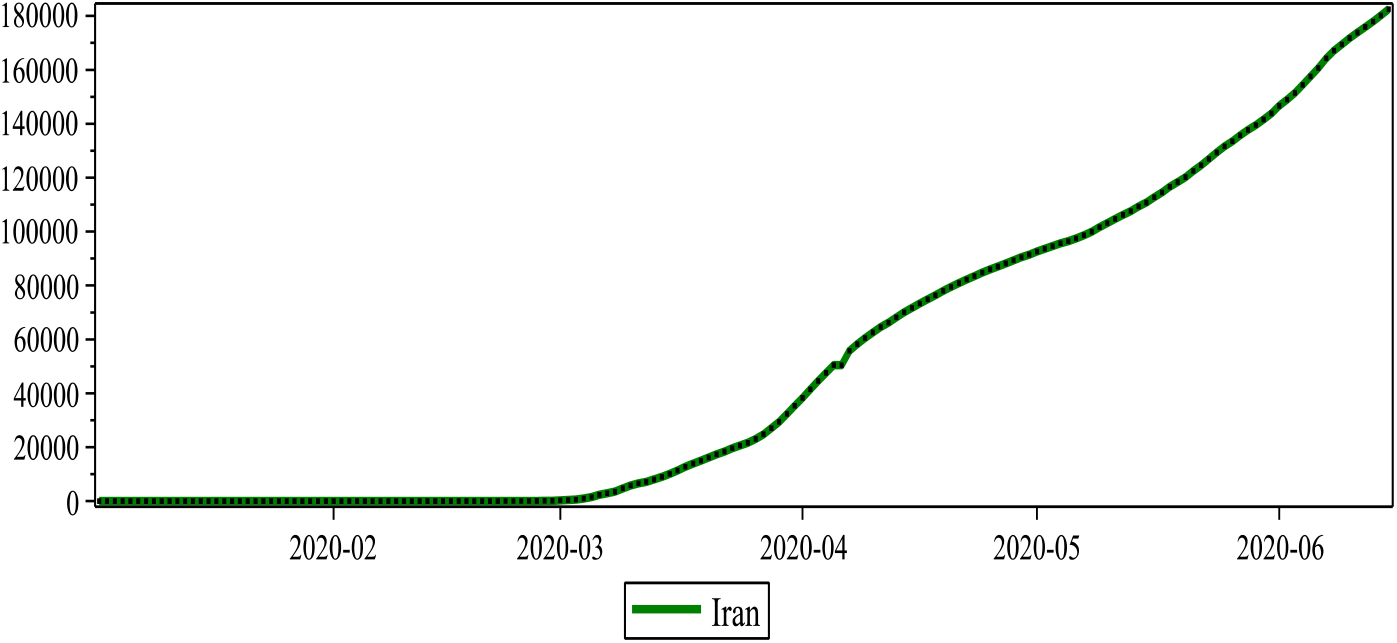
The history of the epidemic in Iran. The green line indicates the daily evolution of total cases.

As evident from the figure above, in mid-April, the second “wave” of the epidemic rose in Iran and at present time the process of spreading the pandemic is far from over.

Let us try to identify the first wave of the epidemic, mathematically evaluating the absolute and relative errors between the input and forecast data, which are described by the logistic curve. The mean squared error is minimal if the first wave reaches the threshold at 93 days from the start of the epidemic, as shown in Fig. 2.

**Fig. 2.**
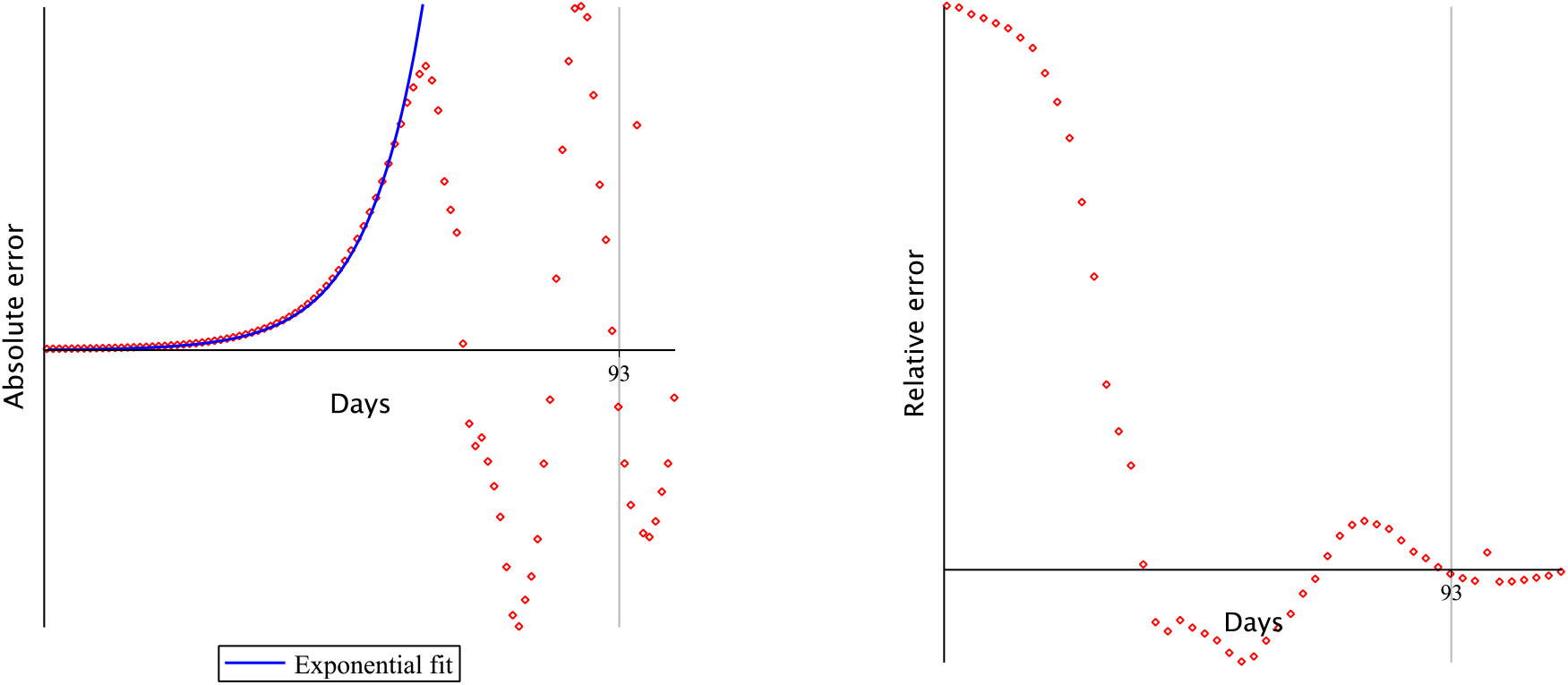
The absolute and relative errors of forecasting the first wave. On the left, the blue line traces the exponential approximation of the discrepancy between actual and forecast data [6].

Thus, the first “wave” is adequately described by a discrete logistic equation with the right-hand side 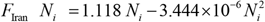. The solution to this equation is shown in Fig. 3.

**Fig. 3.**
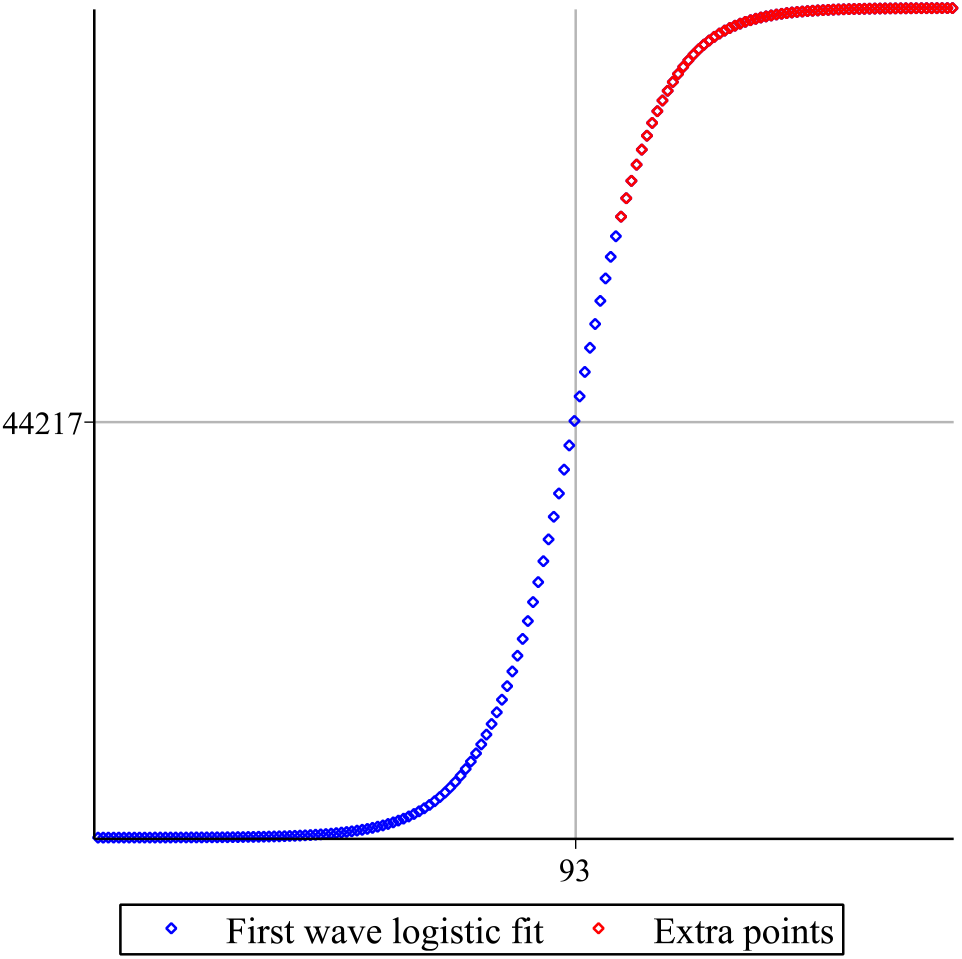
Profile of the first wave of the epidemic in Iran. The red dots display extrapolation points reaching the epidemic peak of the first wave.

Now we decompose the input data taking into account that the logistic curve shown in the above figure adequately approximates the early history of the epidemic. Figure 4 displays this decomposition graphically.

**Fig. 4.**
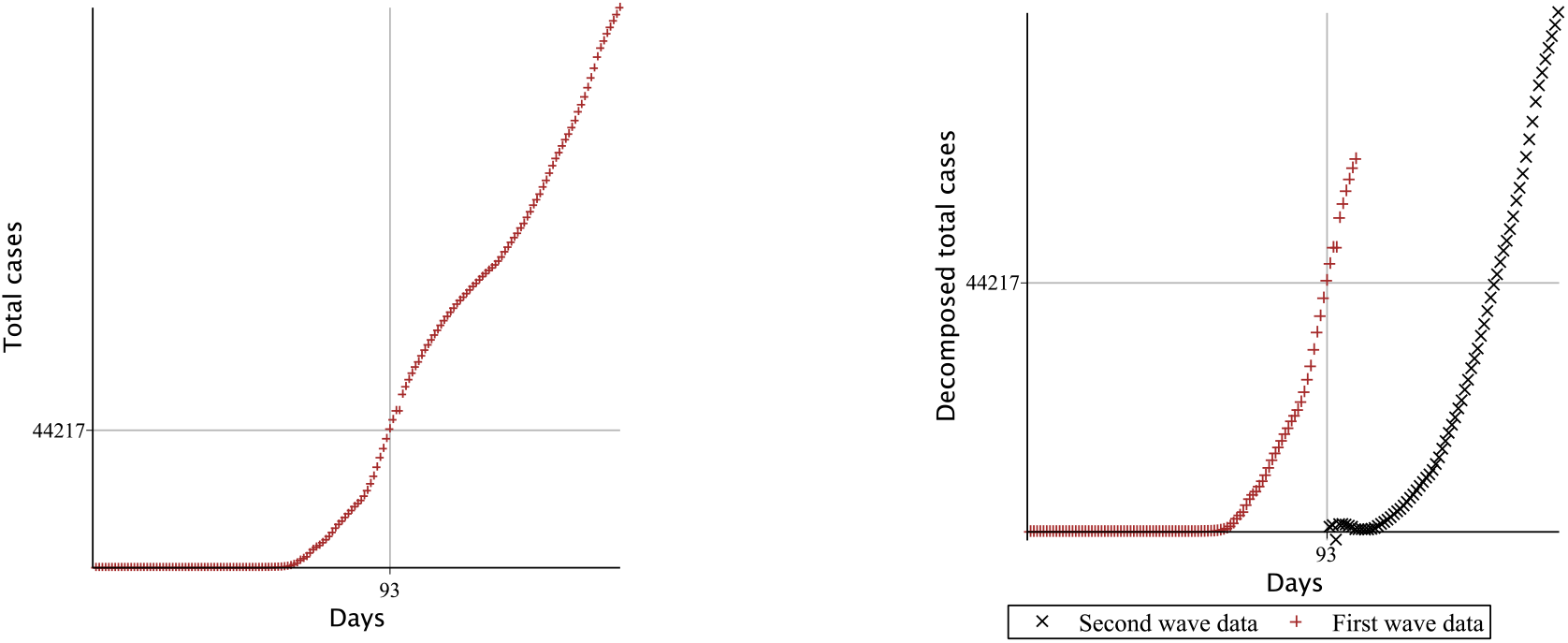
Decomposition of input data. The inset on the left represents the full set of total cases in Iran, for comparison.

At the next stage of processing the input data, the second “wave” of the epidemic is identified. Calculations are performed recursively in the same way as used above. The second “wave” is also adequately described by a discrete logistic equation, but with a different right-hand side 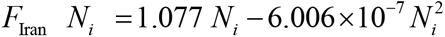. The result is shown in fig. 5.

**Fig. 5.**
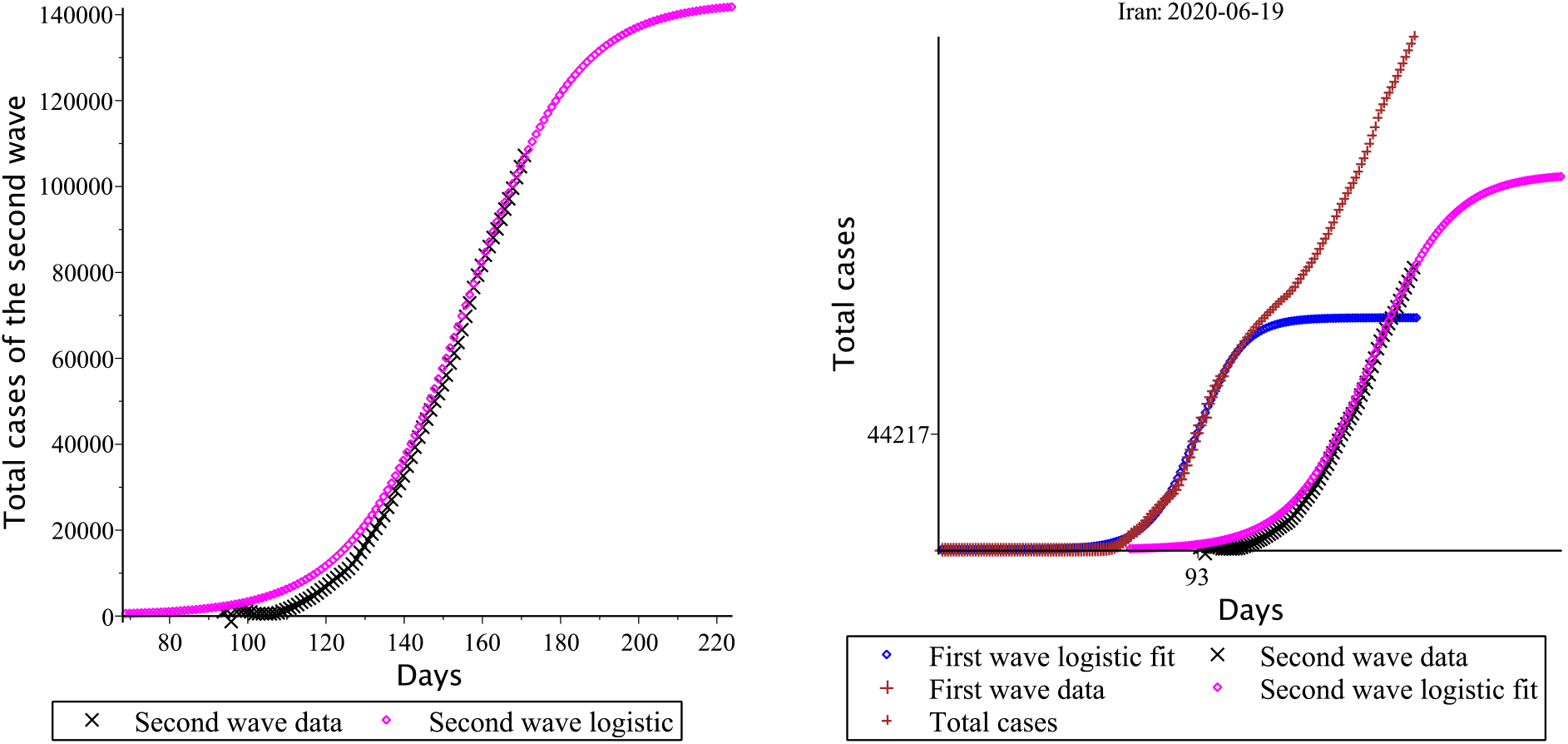
The inset on the left explains the graphical identification of the epidemic second wave in Iran. The right inset shows the assembly of the complete solution to the problem in the form of a superposition of two logistic lines both of the first and second waves.

## Results

The above reasoning and calculations can be repeated using other data on the epidemic history in other countries, for example, Sweden. The history of the Swedish epidemic is shown in fig. 6.

**Fig. 6.**
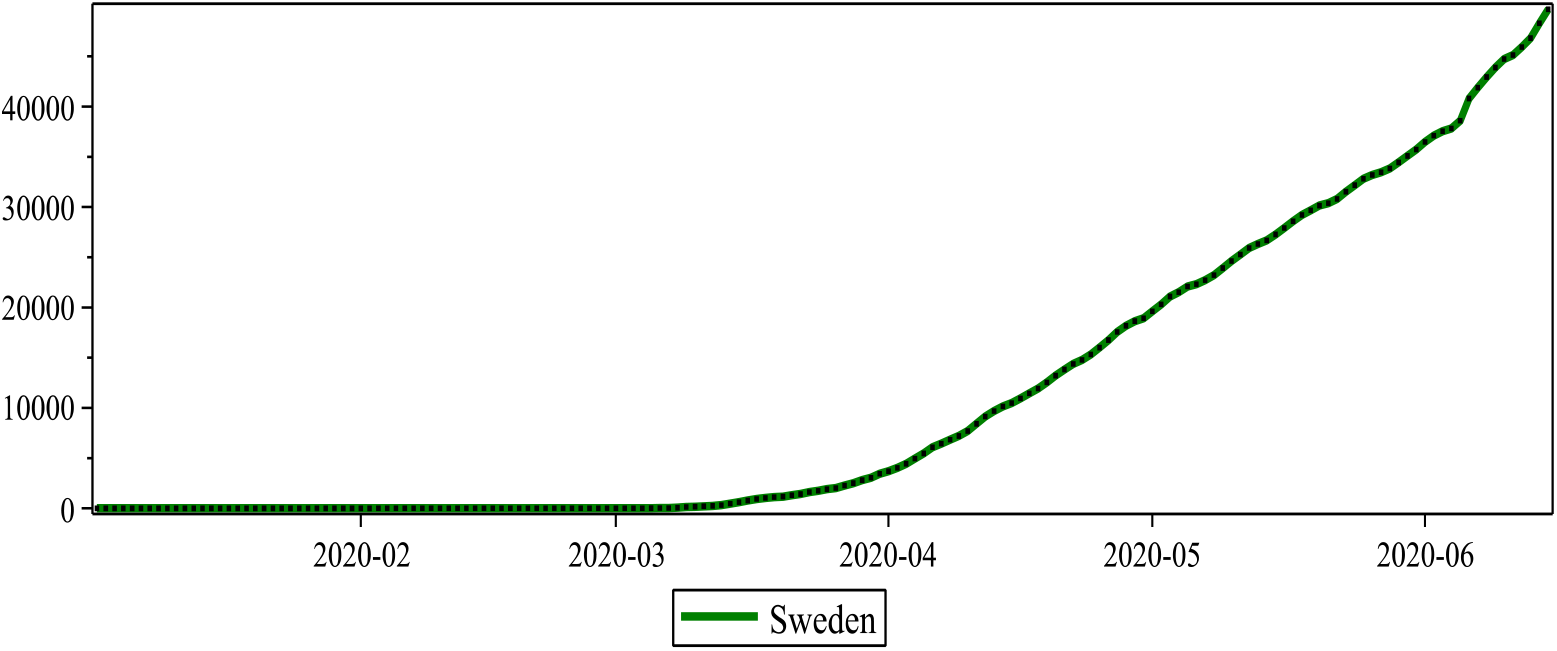
Epidemic history in Sweden. The green line indicates the total daily cases, starting from the very beginning of the epidemic in the country.

The first “wave” of the epidemic in Sweden, compared with Iran, had a relatively small height with a threshold of 7160 recorded cases reached 99 days after the epidemic began in that country, as shown in Fig. 7.

**Fig. 7.**
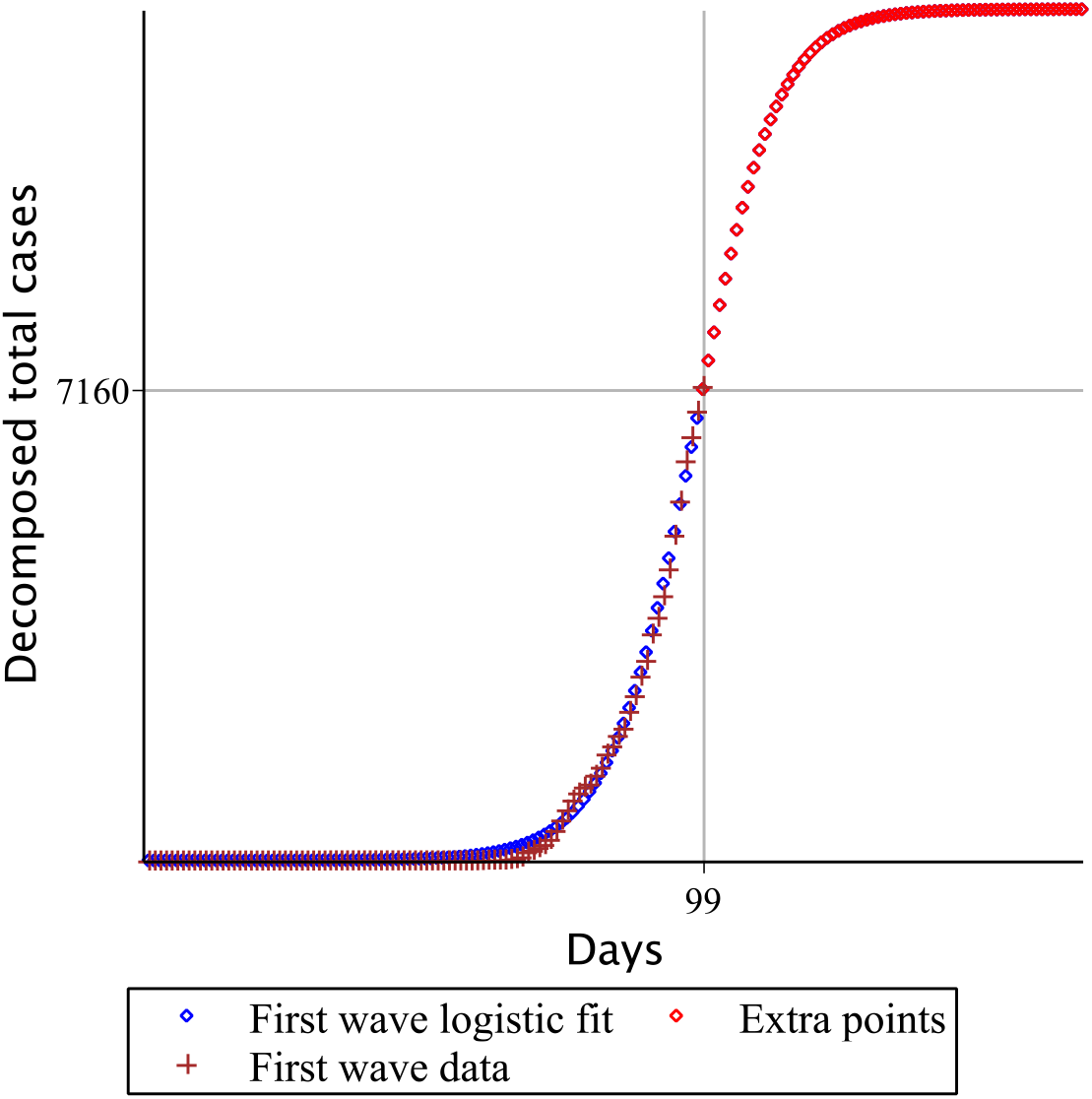
Profile of the epidemic first wave in Sweden. The red dots indicate the extrapolation points reaching the peak of the first epidemic wave.

Decomposition of the input data on the secondary “wave” leads to the result shown in Fig. 8.

**Fig. 8.**
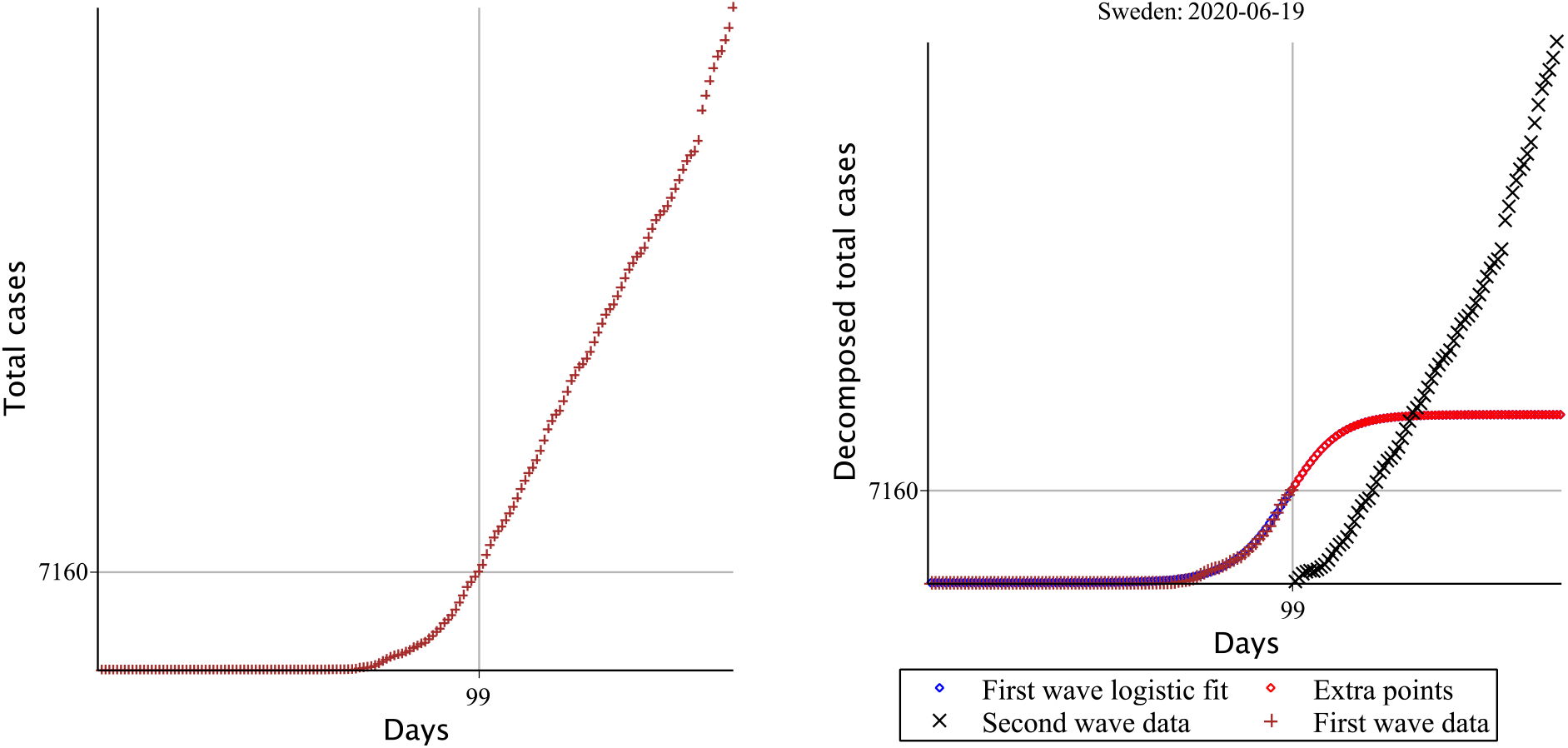
The left inset displays the full set of the input data by total cases in Sweden. The right inset explains the primary decomposition as a superposition of the logistic line of the first wave and the rest related to the secondary waves.

Unlike Iran, the rest of the data corresponding to the secondary waves we cannot already represent as a single logistic line. Thus, the epidemic in Sweden has taken the form of a continuous wave cascade. It is probably the most unfavourable for the combating the epidemic. A situation similar to that observed in Sweden also takes place in the United States.

The rise of the second wave has similar features, despite some differences in the spread of the epidemic in different countries. So, passing the threshold of the current wave should be the most responsible in terms of strict observance of the rules of self-isolation and other sanitary standards. The secondary waves of the epidemic are insidious in that they do not appear immediately but after a sufficiently long time. For example, in Iran, the second wave significantly showed itself in no less than 60 days. To the credit of Iranian specialists, we have admitted that the second epidemic wave in Iran was quickly detected to damp it with maximum efforts. The situation in Sweden seems somewhat neglected. We observe the sequences of secondary epidemic waves with relatively small height, but the dynamics of the process is too sluggish. The situation in the United States is complicated. This case requires additional time and diligence to study.

## Conclusion

This text has focused on the appearance and rise of the second epidemic wave in Iran. An analysis of the input data by the total cases shows that the most favourable time for the emergence of secondary waves is the threshold point in which the rate of the epidemic increase approaches maximum. So, passing the threshold of the current wave should be the most responsible in terms of strict observance of the rules of self-isolation and other sanitary standards.

The algorithm for studying the secondary waves represents a recursive decomposition of the input data to a set of different logistic curves, the parameters of which, unknown in advance, the calculation process determines. At the first step, we check the possibility of adequate approximation of the input data by a single logistic curve. If success, we attempt to identify the second logistic wave, etc.

Logistics decomposition is in many features similar to the well-known Fourier method or wavelet analysis. However, in our case, the decomposition algorithm does not end with a result if the input data is inappropriate.

## Data Availability

I declare the availability of all the data mentioned in the manuscript

https://covid.ourworldindata.org/

